# AUGMENT: a framework for robust assessment of the clinical utility of segmentation algorithms

**DOI:** 10.1101/2024.09.20.24313970

**Authors:** Cathal McCague, Thomas Buddenkotte, Lorena Escudero Sanchez, David Hulse, Roxana Pintican, Leonardo Rundo, AUGMENT study team, James D. Brenton, Dominique-Laurent Couturier, Ozan Öktem, Ramona Woitek, Carola-Bibiane Schönlieb, Evis Sala, Mireia Crispin Ortuzar

## Abstract

**Background:** Evaluating AI-based segmentation models primarily relies on quantitative metrics, but it remains unclear if this approach leads to practical, clinically applicable tools.

**Purpose:** To create a systematic framework for evaluating the performance of segmentation models using clinically relevant criteria.

**Materials and Methods:** We developed the AUGMENT framework (Assessing Utility of seGMENtation Tools), based on a structured classification of main categories of error in segmentation tasks. To evaluate the framework, we assembled a team of 20 clinicians covering a broad range of radiological expertise and analysed the challenging task of segmenting metastatic ovarian cancer using AI. We used three evaluation methods: (i) Dice Similarity Coefficient (DSC), (ii) visual Turing test, assessing 429 segmented disease-sites on 80 CT scans from the Cancer Imaging Atlas), and (iii) AUGMENT framework, where 3 radiologists and the AI-model created segmentations of 784 separate disease sites on 27 CT scans from a multi-institution dataset.

**Results:** The AI model had modest technical performance (DSC=72±19 for the pelvic and ovarian disease, and 64±24 for omental disease), and it failed the visual Turing test. However, the AUGMENT framework revealed that (i) the AI model produced segmentations of the same quality as radiologists (*p*=.46), and (ii) it enabled radiologists to produce human+AI collaborative segmentations of significantly higher quality (*p*=<.001) and in significantly less time (*p*=<.001).

**Conclusion:** Quantitative performance metrics of segmentation algorithms can mask their clinical utility. The AUGMENT framework enables the systematic identification of clinically usable AI-models and highlights the importance of assessing the interaction between AI tools and radiologists.

**Summary statement:** Our framework, called AUGMENT, provides an objective assessment of the clinical utility of segmentation algorithms based on well-established error categories.

**Key results:**

- Combining quantitative metrics with qualitative information on performance from domain experts whose work is impacted by an algorithm’s use is a more accurate, transparent and trustworthy way of appraising an algorithm than using quantitative metrics alone.
- The AUGMENT framework captures clinical utility in terms of segmentation quality and human+AI complementarity even in algorithms with modest technical segmentation performance.
- AUGMENT might have utility during the development and validation process, including in segmentation challenges, for those seeking clinical translation, and to audit model performance after integration into clinical practice.

## 1. Introduction

In the past decade, there has been an explosion of interest in medical devices based on artificial intelligence (AI) and machine learning (ML) technologies. The majority of devices seeking the European Conformité-Européene (CE) mark and Food and Drug Administration (FDA) approval are for radiological use, in part due to the demand for radiological imaging outstripping the number of trained readers, and the proficiency AI/ML-based tools have shown in medical image analysis tasks(1,2). However, despite the increasing number of tools being approved by regulators, their use in actual clinical care remains low, as physician scepticism around their performance and a lack of trust in their black-box nature persists(3). This scepticism is linked to uncertainty about the performance of the tools, and the disconnect between the AI/ML-developers and clinicians who will ultimately be responsible for their use(4).

The most popular area of medical image analysis research relates to segmentation, which denotes the ability to detect a volume of interest and determine its boundaries(5). The performance of segmentation algorithms is quantified by measuring the agreement between the algorithm’s output and the usually human derived “ground truth.” This agreement is expressed using quantitative metrics, with the Dice similarity coefficient (DSC) most commonly used(6). However, there are several issues with this quantitative-metric-only (QMO) based approach to performance assessment.

First, there is an acceptance that such quantitative metrics can be “gamed” to overstate model performance(7).

Second, how performance as measured by the QMO approach translates to real-world practical value has been questioned(5,8). In some instances these metrics may underestimate the clinical utility of an algorithm, and in others they may overestimate it(4). Commonly used metrics such as DSC do not by themselves give qualitative information as to the types of segmentation errors an algorithm is making, instead only measuring the degree of agreement between the algorithm’s segmentation and that of the “ground truth”(9). Research groups developing medical image analysis tools often lack clinical experts, which can lead to basic errors that are obvious to clinicians being made and substantial development time being invested in tools that are ultimately not viable for clinical application(10).

Third, clinician involvement in the QMO evaluation approach is largely passive, often limited only to the generation of the “ground truth” segmentations(4). Clinician inspection of an algorithm’s output is also not commonplace or mandatory, but there is widespread consensus that this should happen routinely and that clinicians should be involved in the development and appraisal of these tools(11). There are, however, currently no clear frameworks on how they should be involved or what form that appraisal should take(12). In addition to this, the current framework of model assessment does not evaluate how well an algorithm collaborates with a clinician, and if its maximal utility may be in an assisting, rather than stand-alone, role.

In this paper we propose a framework for appraising the performance of segmentation models, which assesses practical value in a clinical setting and provides an objective, qualitative evaluation of utility. We chose a challenging segmentation task, namely the segmentation of high grade serous ovarian cancer (HGSOC), for which the state of the art is far from optimal, to demonstrate the added value of qualitative performance assessment with respect to standard metrics. The framework provides added information to AI developers on the types of errors their segmentation model may be producing and fosters closer collaboration with clinical domain-experts during the model development process, thus adding increased transparency and clinical insight to algorithm design.

## 2. Materials and Methods

### 2.1 The AUGMENT framework

We defined and applied a novel qualitative assessment framework called AUGMENT (Assessing Utility of segMENtation Tools) (see Fig. 1). The framework has three steps:

**Figure 1.**
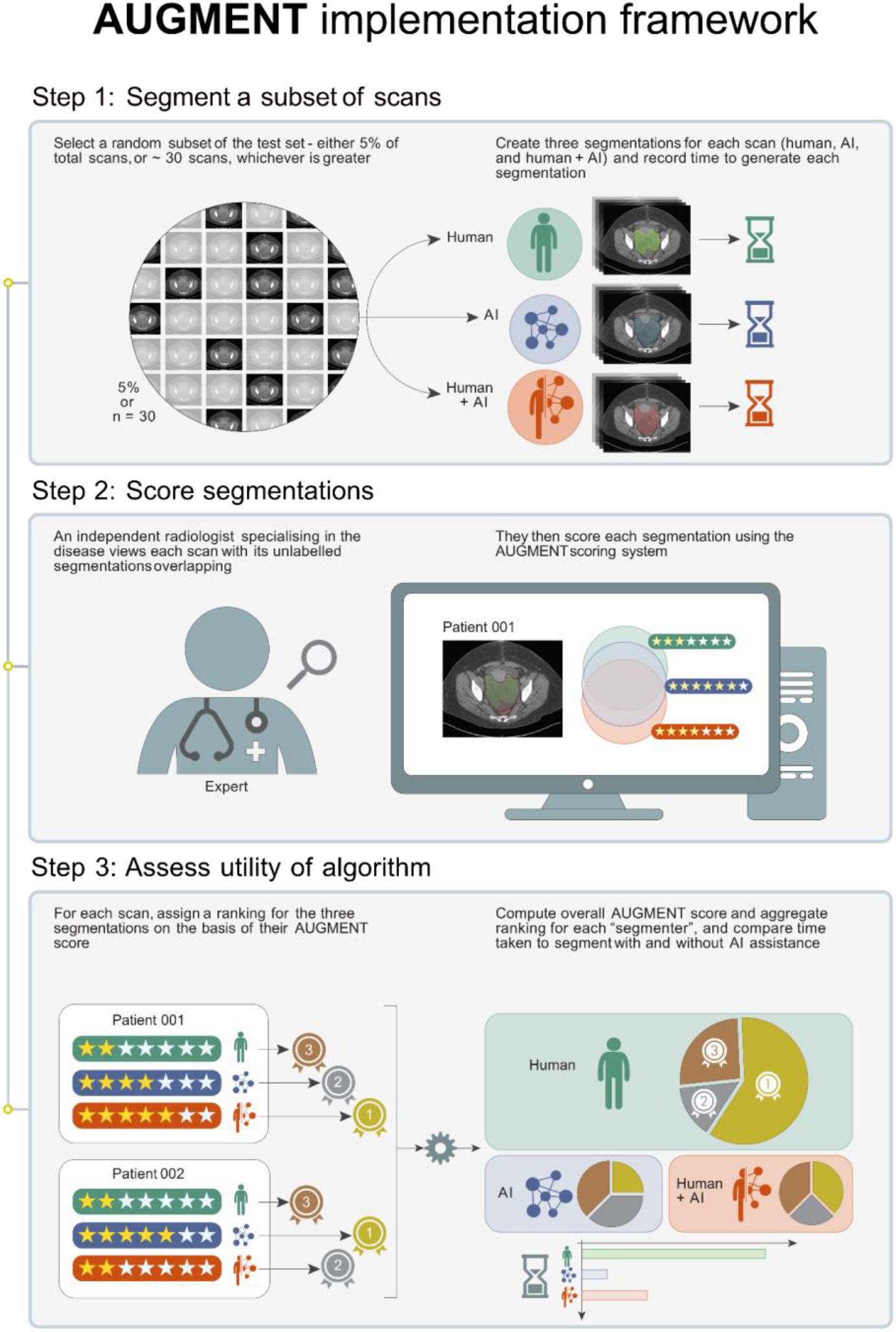
AUGMENT implementation framework, illustrating how the framework can be applied. *Step 1:* A random subset of scans from the validation set (or other dataset, depending on the requirements of the user) are selected at random. Each scan is segmented by all possible segmenter combinations: i) human alone, ii) AI alone and iii) human+AI in collaboration, with time taken to segment also recorded. *Step 2:* An independent clinical domain-expert views each scan with its unlabelled segmentations overlayed on one another and scores each using the AUGMENT scoring system (see Fig. 2). *Step 3:* A ranking of the segmentation approach is made based on the AUGMENT score for each scan. In the event of 2 or more segmentations having equal AUGMENT score the segmentations are given equal ranking (as their clinical utility may later be distinguished by the time-to-segment). Rankings are aggregated to establish the overall ranking of segmentation approaches across the sample-set. The overall AUGMENT score for each segmentation approach (i.e. human, AI, human+AI) is also calculated. The total time for the human to segment the scans alone and with the assistance of the AI is computed to establish if there is a time saving in using the AI as a segmentation assistant.

1. Segmentation of a subset of scans using a human segmenter, an AI segmenter, and a human+AI collaboration;
2. Scoring by an independent domain expert of all unlabelled segmentations using the AUGMENT scoring system (see Fig. 2); and
3. Assessing utility of algorithm by ranking scores and comparing timings.

**Figure 2.**
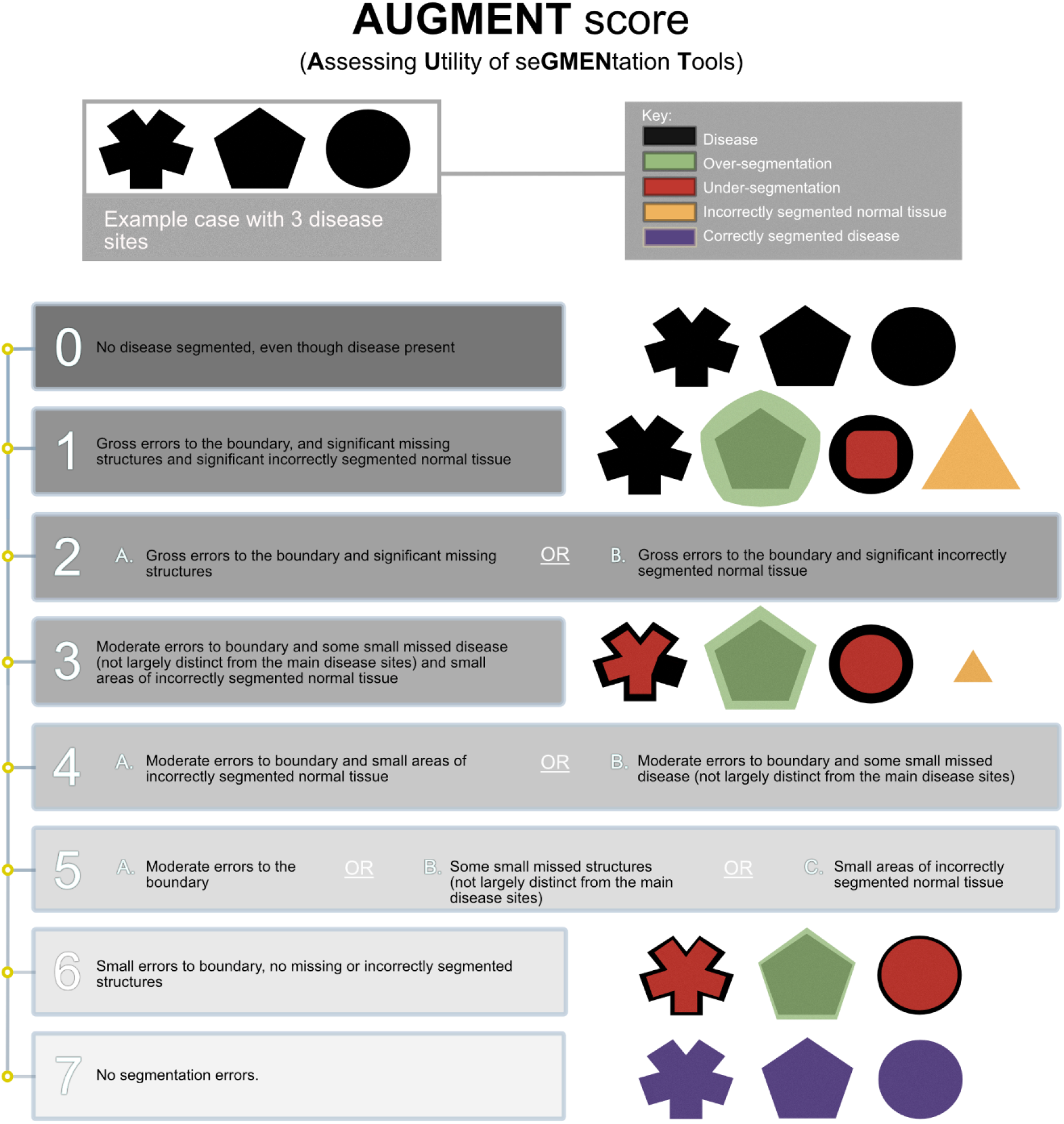
AUGMENT scoring system (with representative visual examples). AUGMENT scores 2, 4 and 5 each contain subcategories. For each of these subcategories their AUGMENT score remains the same, but the inclusion of subcategories are intended to give the developer information on whether the segmentations contain false positive (i.e. incorrectly segmented normal tissue) or false negative (i.e. missed areas of disease) findings.

The AUGMENT scoring system (Fig. 2) has been developed using the main categories of error in segmentation tasks, namely (6,13):

- Quantity, or number of segmented regions
- Area of segmented objects
- Contour of the segmentation
- Content of the segmented region

The scoring system is set up as an 8-level ordinal scale, with 0 being the lowest quality score, and 7 indicating the highest possible quality (no errors identified). The individual scores are explained in more detail in supplementary materials E.

### 2.2 Framework validation

We trained an nnU-net based framework to segment the two most common sites of disease in high-grade serous ovarian cancer patients (HGSOC), namely the omental (OD) and the pelvic (PD) disease sites(14). The model was trained using n=276 scans from 157 patients from an internal dataset, validated using n=104 scans from 53 patients of an external institution and tested on a third independent dataset of n=71 scans from 71 patients from two institutions based in another country, as previously described(14).

### 2.1 Assessing segmentation quality

We used three different frameworks to evaluate the performance of the automatic segmentation algorithm, namely DSC, Visual Turing Test, and a novel framework called AUGMENT.

#### DSC

We calculated the DSC between the automated segmentation and the ground truth, delineated manually by a team of radiologists, with all segmentations reviewed by RW, a board-certified radiologist with 10 years of experience.

#### Visual Turing Test

To assess if the modest DSC scores translated to segmentations noticeably different from manual segmentations we conducted a visual Turing test (VTT) with 20 interrogators, using baseline CT scans of biopsy confirmed ovarian cancer patients from The Cancer Imaging Archive(15). The test contained 80 randomly ordered abdomino-pelvic CT scans, within which were 40 manual (20 PD, 20 OD) and 40 automated (20 PD, 20 OD) segmentations with a total of 429 segmented connected-components. The manual subset was segmented by CM (a specialist trainee in Radiology with 3 years’ experience segmenting ovarian cancer) and checked by RW. In addition to classifying each segmentation as either automated or manual, VTT-interrogators were also asked to make an assessment of the quality of each segmentation, in the form of free-text comment and a 5-point Likert scale score (1 = poorest quality, 5 = greatest quality) without specific guidelines around the meaning of each quality score.

#### Clinical utility assessment using AUGMENT framework

The AUGMENT assessment of the algorithm’s performance was made on a random selection of 27 CT scans acquired during the ICON8 study(16). All were baseline scans acquired at time of diagnosis from patients with biopsy-confirmed high grade serous ovarian carcinoma (HGSOC) who consented to their imaging being used for research purposes after ethical review (REC reference: 20/HRA/2261). For each scan 3 independent segmentations (both PD and OD) were produced by different means:

1. By the model as described above (trained and configured identically to the Turing test configuration).
2. By 3 radiology trainees (9 scans each).
3. By collaborative-segmentation, where a trainee radiologist amended the model’s segmentation (not the same scan as the trainee had previously segmented).

Overall, across the 3 segmentation versions for each scan, there were a total of 784 segmented connected components. The radiologist segmenters in question were CM, DH (specialist trainee in radiology with 6 years experience) and RP (specialist trainee in radiology with 5 years experience).

Following this, an expert genitourinary (GU) radiologist, RW, reviewed the 3 unlabelled segmentations of each scan simultaneously by overlaying the segmentations on top of one another, and gave each segmentation a quality score from 0-7 (0 being of lowest quality, 7 of highest quality) using the AUGMENT scoring tool (see Fig. 2 for scoring chart; additional figure illustrating the types of errors in ovarian cancer can be found in the supplementary materials).

### 2.2 Statistical analysis

For the VTT, a generalised linear mixed model was used to take the potential within-assessor and within-image dependences into account. For the clinical utility assessment we modelled the quality score and time by means of linear mixed models with crossed random effects: random intercepts for patients and segmenters allowing to take the within-patient and within-segmenter dependence into account. All statistical analysis was performed in R version 4.1.2

(2021-11-01) (R Foundation for Statistical Computing, Vienna, Austria. URL https://www.R-project.org/), running on macOS Big Sur 10.16.

## 3. Results

### 3.1 DSC metric

The performance of the model on an external test-set was 72±19 for the PD and 64±24 for the OD lesions. These results have been presented elsewhere(14).

### 3.2 VTT

20 interrogators across the spectrum of expertise were recruited for the VTT. The composition of the interrogator groups is detailed in Table 1.

**Table 1.**
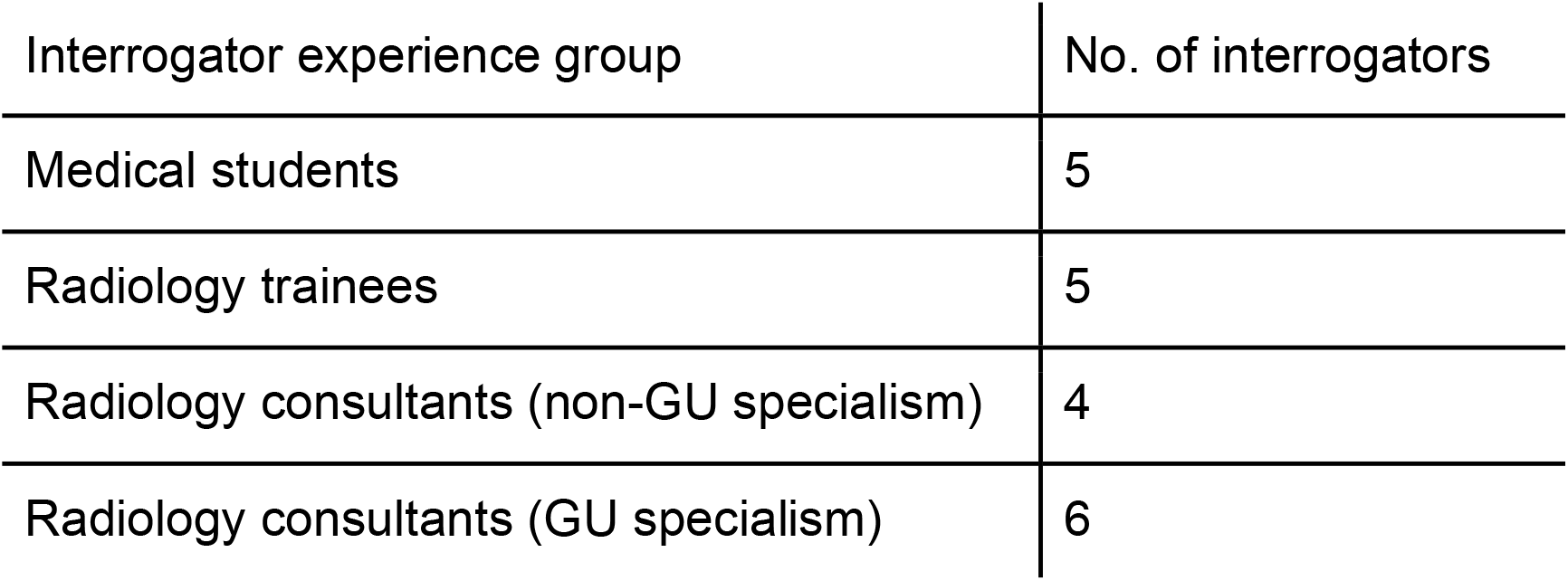
Interrogator groups for VTT.

The overall probability of correctly identifying the origin of the segmentation was significantly greater than the 0.5 score that would signify interrogators were unable to distinguish the AI from human segmentations, therefore the AI failed the Turing test (see Fig. 3 B). However, experience in ovarian cancer imaging, or radiological imaging in general, did not confer an advantage to interrogators (see Table 2): there was no statistical difference between the interrogator groups (Likelihood ratio test *p*=.49) or between PD and OD sites (Wald t-test *p*= .43, see Fig. 3 C). When answer time was assessed (correcting for the experience gained as the interrogators went through the test) there was no difference in time taken to answer the questions between experience groups.

**Table 2.** Classification accuracy for interrogator experience groups.

| Interrogator experience group | Mean classification accuracies |
| --- | --- |
| Medical students | 0.62 |
| Radiology trainees | 0.66 |
| Radiology consultants (non-GU specialism) | 0.60 |
| Radiology consultants (GU specialism) | 0.67 |

**Figure 3.**
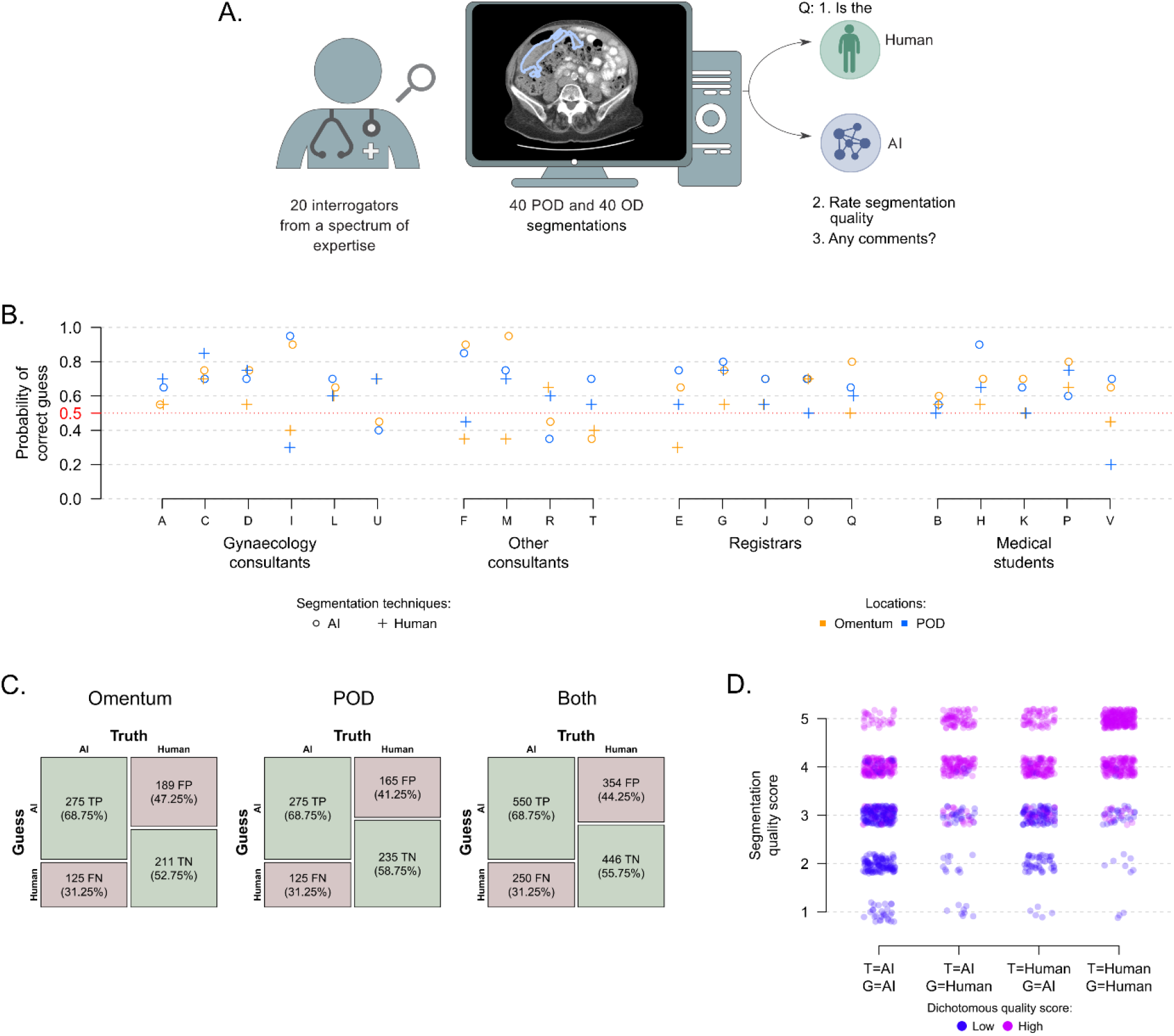
Overview of VTT: A. VTT pipeline. B. Probability of correctly identifying the origin of the segmentation by interrogator. C. Confusion matrix of overall accuracy of interrogator assessment for both VTT (PD and Omentum), and the combined accuracy. D. The figure shows the segmentation quality assessment (y-axis) per combination of actual segmentation technique (T) and guessed segmentation technique (G) levels. Observations are jittered to allow visualisation of the number of observations per category and colour coded by the dichotomous quality assessment measure, which differentiates scores above 3 from scores below 3.

Interrogators from all experience groups noted the difficulty of differentiating between manual and automatic segmentations. The errors they reported fell into 5 broad categories, with examples given in Table 3.

**Table 3.** Interrogator written feedback on the types of segmentation errors noted.

| Types of errors | Example comment |
| --- | --- |
| Segmentations appeared “unnatural” | Interrogator H (medical student): Some segmentations missed low density parts of the tumour leaving a hook / acute angle shape behind. I assumed these would be AI as a human reader would possibly consider such an acute angle “unnatural.” |
| Border errors | Interrogator P (medical student): Generally, the most common error I noticed was that segmentation borders were drawn short of the actual borders of the lesions, leaving some areas of disease unmarked. This was more pronounced with hazy/spiculated borders, where the edges were unclear. |
| Under segmented disease | Interrogator C (genitourinary radiology consultant): Peritoneal thickening/nodularity in lower pelvis appeared to be under segmented (I assume these are the automated rather than the manual analyses). |
| Missed disease | Interrogator O (radiology trainee): There were only a few instances where parts of the pelvic/omental disease were missed and those were difficult to separate from the surrounding normal structures. |
| Incorrectly segmented normal tissue | Interrogator D (genitourinary radiology consultant): Occasional segmenting of the bowel if there was no gas in the lumen. |

On average, lowest scores were given to images correctly identified as being segmented by AI, and highest scores were given to images correctly identified as being segmented by humans (see Fig. 3 D). Interrogators gave higher quality scores to AI segmentations they thought were human (Truth: AI, Guess: human), and lower scores to human segmentations which they thought were AI (Truth: human, Guess: AI) which suggests there was some bias towards segmentations they perceived as produced by the AI.

### 3.3 Clinical utility assessment using AUGMENT framework

The average AUGMENT score for human segmentations was 3.22. The difference between AI segmentation AUGMENT score and the human segmentation AUGMENT score was not significant (*p*=.46). The difference between human+AI segmentation AUGMENT score and the human segmentation AUGMENT score was significant (t-test, *p*=<.001), with an average AUGMENT score for this group of 4.59.

The segmentation rankings per scan (based on the AUGMENT score) were: human+AI segmentations = 1st place (or joint 1st) in 19/27 scans (70%); AI segmentations = 1st place (or joint 1st) in 11/27 scans (41%); human segmentations = 1st place (or joint 1st) in 6/27 scans (22%) (see Fig. 4 B).

**Figure 4.**
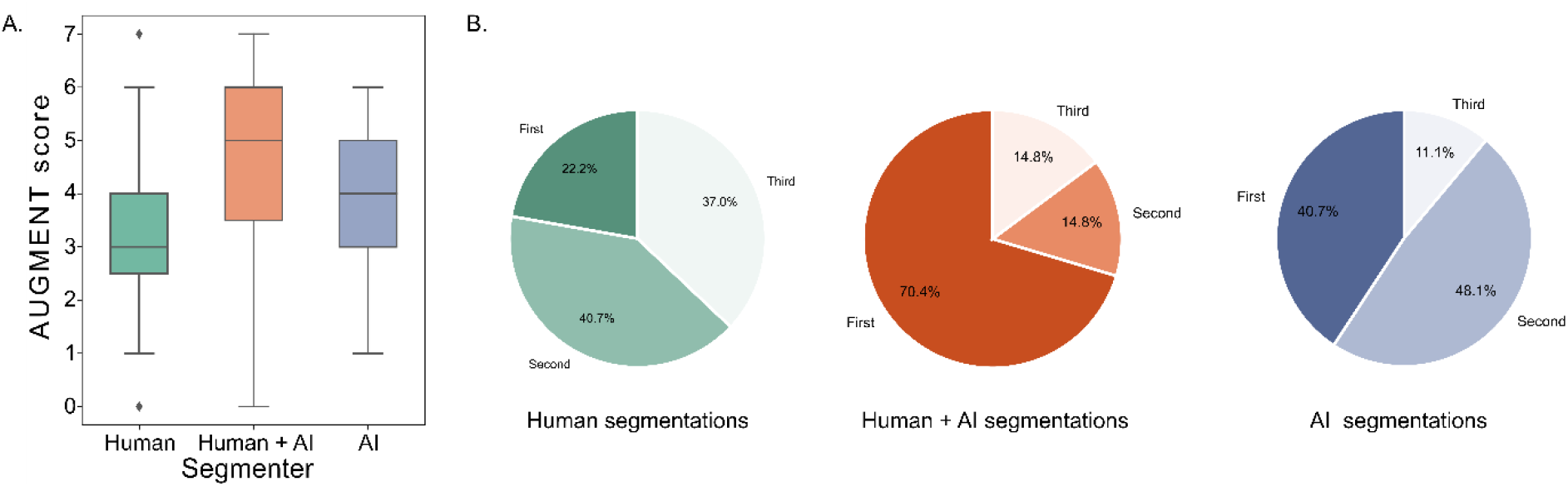
A. Boxplots of the AUGMENT score by segmenter, showing minimum, maximum, 1st quartile and 3rd quartile. B. Pie charts of the rankings for each segmentation mode.

There was a time-saving effect for the combined human+AI method compared to the baseline human segmentation method (t-test *p*=<.001) leading to an average decrease in segmentation time, compared to the baseline, of 57.23% (CI [42.3%, 68.29%]). This means, for example, that a segmentation which takes a radiologist 10 minutes would translate to a segmentation time of ∼ 4 minutes with the help of the AI.

When considering segmentation time by location, there was no statistically significant difference between the human segmentation times of PD and OD at the 5% level (t-test *p*=.22). We compared the additive effect of the AI on segmentation times of the OD and PD, using the OD time as the baseline. This effect is significant (t-test *p*=.02) leading to an average decrease in segmentation time, compared to the baseline, of 47.84% (CI [9.99%, 69.77%]). This means the segmentation time reduction allowed by AI in PD is significantly larger than the gain in segmentation time reduction allowed by AI in OD. Nevertheless, combining human+AI also resulted in significantly shorter segmentation times for OD by itself (t-test p=.01, 40.31% reduction).

## 4. Discussion

We have presented a novel framework to assess clinical utility of segmentation algorithms and compared it to other common evaluation methods in the context of ovarian cancer segmentation. The VTT showed no statistically significant differences in terms of classification accuracy and speed between experience groups ranging from students with no radiological experience to consultants with over a decade sub-specialty experience in genitourinary imaging. Written feedback on the segmentations, as detailed in Table 3, noted no major systematic errors, but some interrogators highlighted occasional unnatural features which, in certain instances, made them suspect creation by the algorithm. These results suggested that there may be clinical utility which is not being captured by the DSC metric assessment of performance.

The AUGMENT evaluation showed that even with modest DSC performance the algorithm did have utility in i) augmenting the segmentation ability of the human radiologists and ii) drastically reducing the segmentation time. The AI segmentations had a higher mean AUGMENT score than those created by the radiologists alone, however this difference was not statistically significant.

The AUGMENT framework can be applied beyond the specific scenario of ovarian cancer as the categories of errors which underpin it are universal to all segmentation tasks(6). Its implementation, in a subset of scans during the testing and validation process, is intended to complement the insights of one or more appropriately chosen quantitative metric(s), not replace their use.

There is cross-discipline agreement that combining quantitative metrics with qualitative information on performance from domain experts whose work is impacted by an algorithm’s use is a more accurate, transparent and trustworthy way of appraising an algorithm than using quantitative metrics alone(7,17,18). A limitation of quantitative metrics is the uncertainty around the ground truth, which in many radiological imaging modalities can be large(8,19). In step 2 of the AUGMENT framework (see Fig. 1) the premise of showing the 3 segmentations one-on-top-of-the-other for direct comparison is underpinned by Structure Mapping Theory(20,21). Such comparisons are known to foster insight in the observer, as they sharpen focus on which commonalities and differences are salient, and invite new inferences not initially obvious(20). Comparing the segmentations simultaneously augments the ability of the independent expert to infer the “ground truth” more closely, allowing them to make a better assessment of the relative quality of each segmentation.

By comparing all permutations of segmentation approach AUGMENT allows for a situation where an algorithm’s maximal utility may be as an assistant to a human. Evaluating how an AI model collaborates with a human, in addition to how it functions alone, is one of the key guiding principles recommended by all major healthcare regulatory agencies(22). In the middle of the scale AUGMENT prioritises a model’s sensitivity, valuing the ability to detect disease that may otherwise be overlooked by the segmenting radiologist over the avoidance of false positives as these mistakes are easily recognised and rectified by a radiologist. Scores 3, 4, and 5 include the clause “not largely distinct from the main disease sites” for small areas of missed disease; this clause is not included for areas of incorrectly segmented normal tissue. This weighting aims to confirm if the AI tool effectively supports radiologists by favouring the detection of disease over falsely identifying normal tissue as pathological.

Our study and the proposed framework have several limitations. First, application of AUGMENT requires the involvement of at least 2 radiologists to perform the roles of i) segmenter(s) and ii) independent expert. This necessitates access to experts, and commitment of their time, however the inclusion of this expertise in the model development team would likely add value to the model under development. Second, the application of AUGMENT may be affected by inter-reader variability. This variability is likely to be mainly focused on the boundary of disease, rather than its presence, with those regions of well defined disease with clear background discrimination presenting less disagreement than those with diffuse or spiculated borders poorly discriminated from the background(23). Boundary disagreements may be heavily penalised by metrics like DSC but represent subjective assessment by individual clinicians which are not clinically impactful. Small inter-reader variances limited to the boundary of disease would not severely impact the AUGMENT score (likely only resulting in downgrading from a score of 7 to a score of 6). Our use-case example used 3 separate segmenters to test the AI’s utility for a variety of segmentation approaches, however the statistical model employed allowed us to take within-patient and within-segmenter dependence into account, and results were strongly significant even with these variances. Third, AUGMENT segmenters were trainee radiologists, however all were experienced ovarian cancer segmenters, with DH and RP at the end of their training having completed fellowship examination. The independent expert, RW, was an experienced radiologist, but the difference in radiology experience within the segmenter group, and between segmenters and independent expert, is of the same order.

The relative ease of entry to the medical image analysis field means those producing both research and commercial tools may be of varying expertise(24). There are many opportunities to introduce errors to algorithm design, which might have downstream consequences. For those final tools which seek full integration in the clinical workflow there will be increased scrutiny of performance. AUGMENT could act as a framework for such local pre-implementation checks, or the continual auditing process.

To implement AUGMENT in the challenge setting, we suggest independent model performance is compared against i) a human alone, and ii) a combined segmentation team of human+model, so that all potential permutations of application are considered. This might result in separate categories of competition, for instance i) best independent segmentation approach, ii) best collaborative segmentation approach, and iii) best overall segmentation approach. Alternatively, the AUGMENT framework could also be applied to compare segmentation models side-by-side without considering the models performance as segmentation aids.

In conclusion, evaluating segmentation model performance purely on the basis of quantitative metrics may be an inaccurate way of capturing quality and utility(5,7). Using a framework like AUGMENT, in tandem with an appropriate quantitative metric, could improve understanding of performance. It could add transparency to the evaluation process by ensuring that clinical domain-experts, whose work is affected by an algorithm’s use, are part of their design. In addition to having potential application during the development and validation process, including in segmentation challenges, AUGMENT might have utility for clinical translation and to audit model performance after integration into clinical practice.

## Data Availability

All data produced in the present study are available upon reasonable request to the authors.

## Ethical approval

This retrospective image analysis study was given ethical approval through the Integrated Image Analysis in High Grade Serous Ovarian Cancer study (IIA-HGSOC) (REC reference: 20/HRA/2261).

Ethical approval for developing and training the AI model used in this study is detailed in (14). The images used in the VTT, from The Cancer Imaging Archive, are used under a Creative Commons Attribution 4.0 International License (15). The images used in the comparison study are from the ICON8 study, the use of which was approved as part of IIA-HGSOC. At the time of the ICON8 study these patients consented to their images being used for further research, and re-consenting for IIA-HGSOC was waived by the research governance and ethics committee.

## Data availability

The code for the segmentation model evaluated in this study is available at https://github.com/ThomasBudd/ovseg/tree/abee8ef38838ffa34f3553624eef3f626ba914bd. The model was trained using CT scans from 3 distinct non-overlapping datasets of biopsy confirmed ovarian cancer patients acquired in the US and UK, which are not currently public (14).

VTT - the images used in the VTT are from biopsy confirmed ovarian cancer patients from The Cancer Imaging Archive, which are publicly available at https://www.cancerimagingarchive.net/. The segmentations used are not currently publicly available.

Comparison study – the images used in comparison study were of randomly selected biopsy confirmed ovarian cancer patients from 2 centres in the UK who participated in the ICON8 study. These images are not currently public.

