## Supplemental materials for "AUGMENT: a framework for robust assessment of the clinical utility of segmentation algorithms"

### Supplementary Materials

#### A. Training the segmentation model:

Neural Network Training. Training was done by applying progressive learning, e.g. by simultaneously increasing input resolution, patch size and augmentation magnitude. Therefore the training was subdivided into four stages with patch sizes and downsampling factors given in Table S1 and magnitude of grey value augmentations given in Table S2. All other augmentations, namely flipping and spatial scaling and rotation were applied as suggested by the authors of nnU-net. The application probabilities of the grey value augmentations were also left unchanged. The application of progressive learning decreased the training time by roughly 40%.

| Stage | Patch size | Downsampling factor per axis |
|---|---|---|
| 1 | 128 x 128 x 40 | 1.688 x 1.688 x 1.6 |
| 2 | 152 x 152 x 44 | 1.421 x 1.421 x 1.455 |
| 3 | 192 x 192 x 60 | 1.125 x 1.125 x 1.067 |
| 4 | 216 x 216 x 64 | 1 x 1 x 1 |

**Table S1.** Input patch sizes and downsampling factors used in the four stages of progressive learning. The downsampling provides that the visual context of the network stays constant during the training.

| Transformation | Stage 1 | Stage 4 |
|---|---|---|
| Additive Gaussian noise | U[0, 0.025] | U[0, 0.1] |
| Gaussian blurring | U[0.125, 0.75] | U[0.5, 1.5] |
| Multiplicative brightness | U[0.925, 1.075] | U[0.7, 1.3] |
| Contrast | U[0.9125, 1.125] | U[0.65, 1.5] |
| Low-resolution simulation | U[1, 1.25] | U[1, 2] |
| Gamma | U[0.925, 1.125] | U[0.7, 1.5] |

**Table S2.** Adaptation of grey value augmentations in progressive learning. Here U[a, b] denotes sampling from a uniform distribution with minimum a and maximum b. Both a and b are linearly increased from stage one to four.

#### B. Creating the automated segmentations

A single model was trained for both PD and OD in a five-fold cross validation scheme using the training data described above. Hyper-parameters were tuned by using a custom

reimplementation of the well established nnU-Net framework (25) aiming to increase the DSC for both lesions sites on the first evaluation set. As a pre-processing step, all images were resized to 0.8 mm in plane pixel spacing and 2.5mm slice thickness, in addition to windowing and normalization of the grey values. To increase the capacity of the network, the encoding path of the U-Net was replaced with a ResNet (26), with one, two, six and three blocks per stage. The network was trained for 250,000 batches of size two using a stochastic gradient descent algorithm with Nesterovs momentum (27). The sum of cross entropy and soft dice loss was introduced to all four stages of the U-Net and a weighted sum as the final loss function was used for the training. The learning rate schedule suggested by the nnU-Net framework was replaced with a linearly increasing warm-up over 5% of the batches followed by a cosine decay (28). During training extensive data augmentation including rotations, scaling, flipping, and various grey value manipulations such as adding Gaussian noise, blurring, and changing contrast were applied. To decrease the computational cost of the training, progressive learning was employed (29). After applying the sliding window algorithm to evaluate the networks on full 3d volumes, ensembling was applied by computing the mean of the softmax outputs of the five networks trained in the cross validation. The implementation of the deep learning model was performed in PyTorch 1.9. During a visual inspection of the models performance on the second evaluation dataset, some artefacts in the automated predictions, namely small connected components, holes in the segmentations, thin lines, and pouches of single pixel width as a part of larger regions of interest, were discovered. As a result, an additional post processing step was introduced for each axial slice including removal of small connected components with less than 10 pixels, filling holes, and binary opening and closing using a 3x3 cross as a structural element, none of which had an impact on the DSC of the predictions. False positive annotations of omental disease in the breast of some scans were also discovered. To address this a U-Net for liver segmentation was trained on the LiTS challenge dataset to identify the axial slice containing the most liver tissue, which was typically close to the diaphragm, and this was used to determine the upper boundary of the segmented region of interest; across the training and evaluation data, only 0.03% of the whole omental disease volume, and no pelvic/ovarian disease, was found above this point.
After freezing all hyper-parameters including the post processing, the model was evaluated on the test dataset, and the predictions were exported to the DICOM format using the rt-utils library. To ensure that no bugs produced obvious errors like upside down flipping or 90 degrees rotations, the automated predictions were visually inspected before releasing the test to the interrogators. Importantly, however, no scans were exchanged or modified, and the segmentations were frozen at this point.

#### C. VTT platform

A completely new platform, based on the OHIF viewer plugin to XNAT[ohifplugin] was developed to fulfil the requirements of the visual Turing test conducted in this paper. An initial Rapid Reader prototype had previously been developed but the new software introduced features specifically tailored to:
i) provide a seamless workflow with a list of cases randomly ordered per user (reader);
ii) allow for “one-click” loading of the pre-existing segmentations (either manually or automatically created);
iii) present the reader with a custom-made electronic case report form (eCRF); and
iv) record the answers from the reader, subsequently moving to the following case in the worklist.

The eCRFs contain the following fields, which are stored in XNAT as assessors and read later for analysis using the XNATPy[xnatpy] Python interface to XNAT:
a) *Assessment* (a drop-down menu allowing the options ‘MANUAL’ or ‘AUTOMATED’);
b) *Confidence* (of the user on chosen assessment, with a drop-down menu allowing as options integers between 1 and 5);
c) *Quality* (reader’s opinion on the quality of the segmentation, with a drop-down menu allowing as options integers between 1; and
5) and
*d) Comments* (free text box).
Additional metadata is created in the assessor with information on the XNAT imaging session ID, creation date and reader ID. A screenshot of the platform is presented in supplementary materials D. The built plugins can be made available upon reasonable request to the authors L.E.S., S.J.D., M.A., J.D., W.C.

**D. Screenshot of the VTT platform**
Screenshot of the platform developed to perform the visual Turing test, based on the OHIF viewer and Rapid Reader plugins to XNAT, showing one case rendered in the OHIF viewer plugin and the custom eCRF panel on the right-hand side.

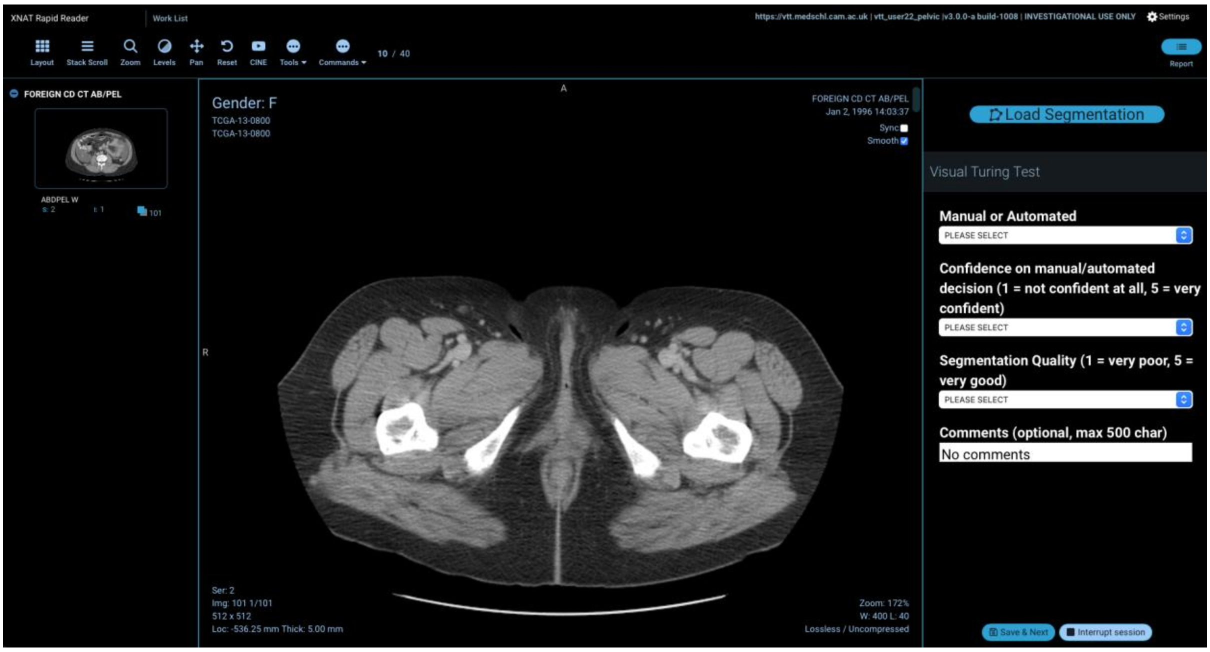


**E. Individual AUGMENT scores explained**
The scores are ordered based on the severity of the errors present, with 0 having most errors and 7 having no errors. AUGMENT scores 2, 4 and 5 each contain subcategories.

- Score 0 - in this case either i) no disease is segmented, even though disease is present, or ii) disease is segmented, but no disease is present. In such cases, all the above errors are considered applicable, and the level of error is most significant. Therefore, this is the lowest extreme of the scale. Note - in the current study Ovseg has only been used to segment scans of biopsy confirmed ovarian cancer, therefore only case i) is possible. However, to make the scoring system applicable for other segmentation tasks which may also assess performance on "normal" disease free scans, category ii) is also included.

- Score 1 - in this case errors 1, 2, 3, and 4, are present, and the level of these errors is significant and wide-ranging. There are gross errors to the boundary, and significant missing structures and incorrectly segmented normal tissue.
- Score 2 - in this case errors 1, 2, 3, and 4, are present, and the level of these errors is significant. There are gross errors to the boundary present. Additionally, either significant missing areas of disease (Score 2 A), or significantly incorrectly segmented normal tissue is present (Score 2 B). For the purposes of aggregating the scores both subcategories A & B have the same AUGMENT score (i.e. 2) but labelling them as either A or B is intended to give the developer information on whether the segmentations contain false negative (i.e. A - missed disease) or false positive (i.e. B - incorrectly segmented normal tissue) findings.
- Score 3 - in this case errors 2, 3, 4, and potentially 1 are present, and the level of these errors is moderate, and wide-ranging. There are moderate errors to the boundary present. Additionally, there are small areas of missing disease (which are not largely distinct from the main disease sites), and there are small areas of incorrectly segmented normal tissue.
- Score 4 - in this case errors 2, 3, and 4, are present, and the level of these errors is small to moderate, and widespread. There are moderate errors to the boundary present. Additionally, there are small areas of missing areas of disease (which are not largely distinct from the main disease sites) (Score 4 A), or small areas of incorrectly segmented normal tissue (Score 4 B). As above, both subcategories A & B have the same AUGMENT score (i.e. 4) but labelling them as either A or B is intended to give the developer information on whether the segmentations contain false negative (i.e. A - missed disease) or false positive (i.e. B - incorrectly segmented normal tissue) findings.
- Score 5 - in this case errors 2, 3, and potentially 4, are present, the level of these errors is small to moderate, and they are not widespread. There is either i) small areas of missing disease (which are not largely distinct from the main disease sites) (Score 5 A), ii) small areas of incorrectly segmented normal tissue (Score 5 B), or iii) moderate errors to the boundary (Score 5 C). This time there are 3 subcategories A, B, & C, which have the same AUGMENT score (i.e. 5). As previously, A denotes false negatives (missed disease), B denotes false positives (incorrectly segmented normal tissue). In addition, C denotes moderate errors in the boundary delineation (these do not fall neatly into the binary classification of false positive or false negative but refer to inaccuracies in precisely outlining the area of interest).
- Score 6 - in this case error 2, 3 and 4 are present, the level of these errors is small, and they are not widespread. There are small errors to the boundary, but no areas of missing disease or incorrectly segmented normal tissue.
- Score 7 - none of the 4 error categories above apply as no segmentation errors are present. Therefore, this is the highest extreme of the scale.

**F. Augment Scoring chart illustrating the types of errors in ovarian cancer**

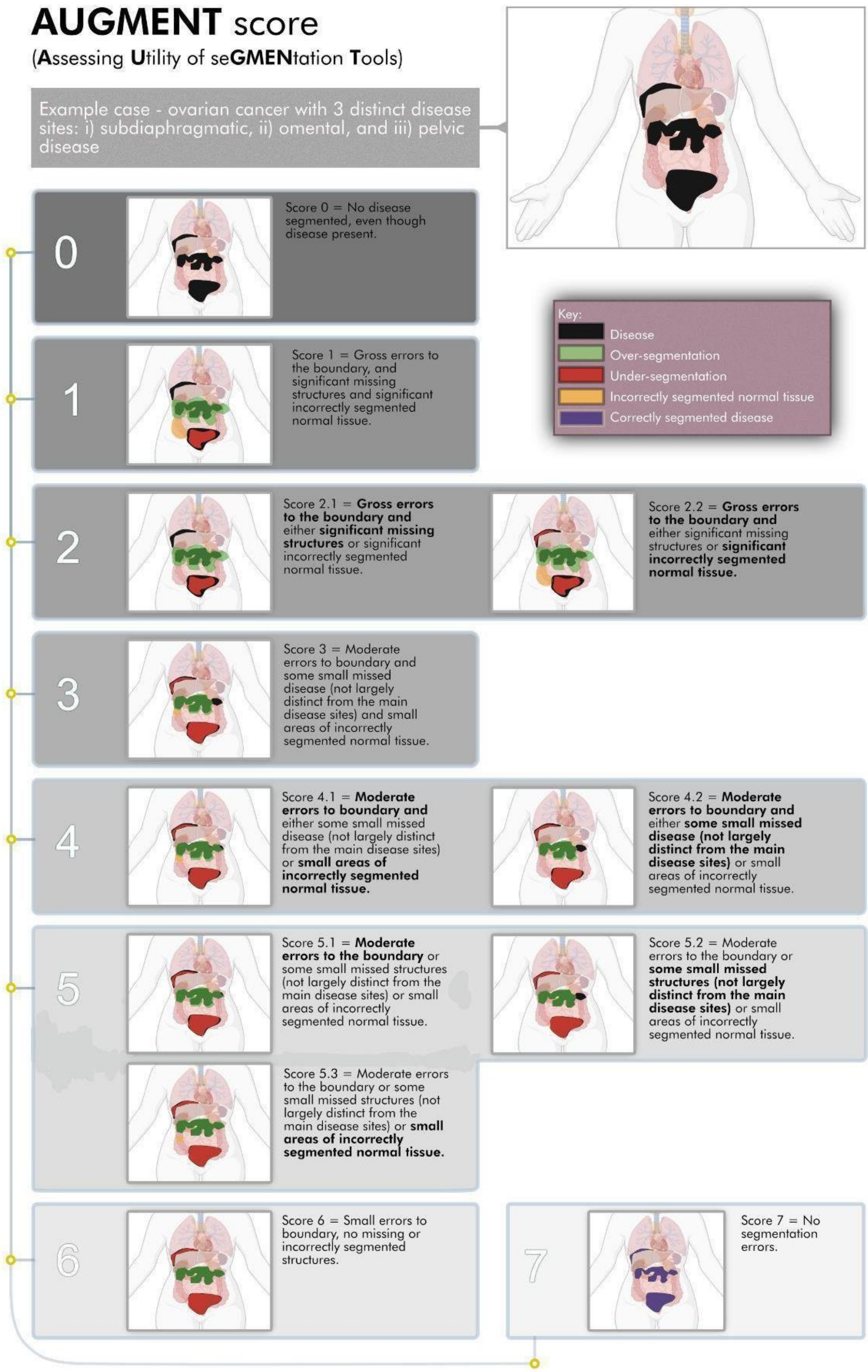

AUGMENT score
(Assessing Utility of seGMENtation Tools)
Example case - ovarian cancer with 3 distinct disease sites: i) subdiaphragmatic, ii) omental, and iii) pelvic disease
Key:
Disease
Over-segmentation
Under-segmentation
Incorrectly segmented normal tissue
Correctly segmented disease
0
Score 0 = No disease segmented, even though disease present.
1
Score 1 = Gross errors to the boundary, and significant missing structures and significant incorrectly segmented normal tissue.
2
Score 2.1 = Gross errors to the boundary and either significant missing structures or significant incorrectly segmented normal tissue.
Score 2.2 = Gross errors to the boundary and either significant missing structures or significant incorrectly segmented normal tissue.
3
Score 3 = Moderate errors to boundary and some small missed disease (not largely distinct from the main disease sites) and small areas of incorrectly segmented normal tissue.
4
Score 4.1 = Moderate errors to boundary and either some small missed disease (not largely distinct from the main disease sites) or small areas of incorrectly segmented normal tissue.
Score 4.2 = Moderate errors to boundary and either some small missed disease (not largely distinct from the main disease sites) or small areas of incorrectly segmented normal tissue.
5
Score 5.1 = Moderate errors to the boundary or some small missed structures (not largely distinct from the main disease sites) or small areas of incorrectly segmented normal tissue.
Score 5.2 = Moderate errors to the boundary or some small missed structures (not largely distinct from the main disease sites) or small areas of incorrectly segmented normal tissue.
Score 5.3 = Moderate errors to the boundary or some small missed structures (not largely distinct from the main disease sites) or small areas of incorrectly segmented normal tissue.
6
Score 6 = Small errors to boundary, no missing or incorrectly segmented structures.
7
Score 7 = No segmentation errors.

### G. Confusion matrices of accuracy of interrogator assessment for both VTT (PD and OD) by assessor

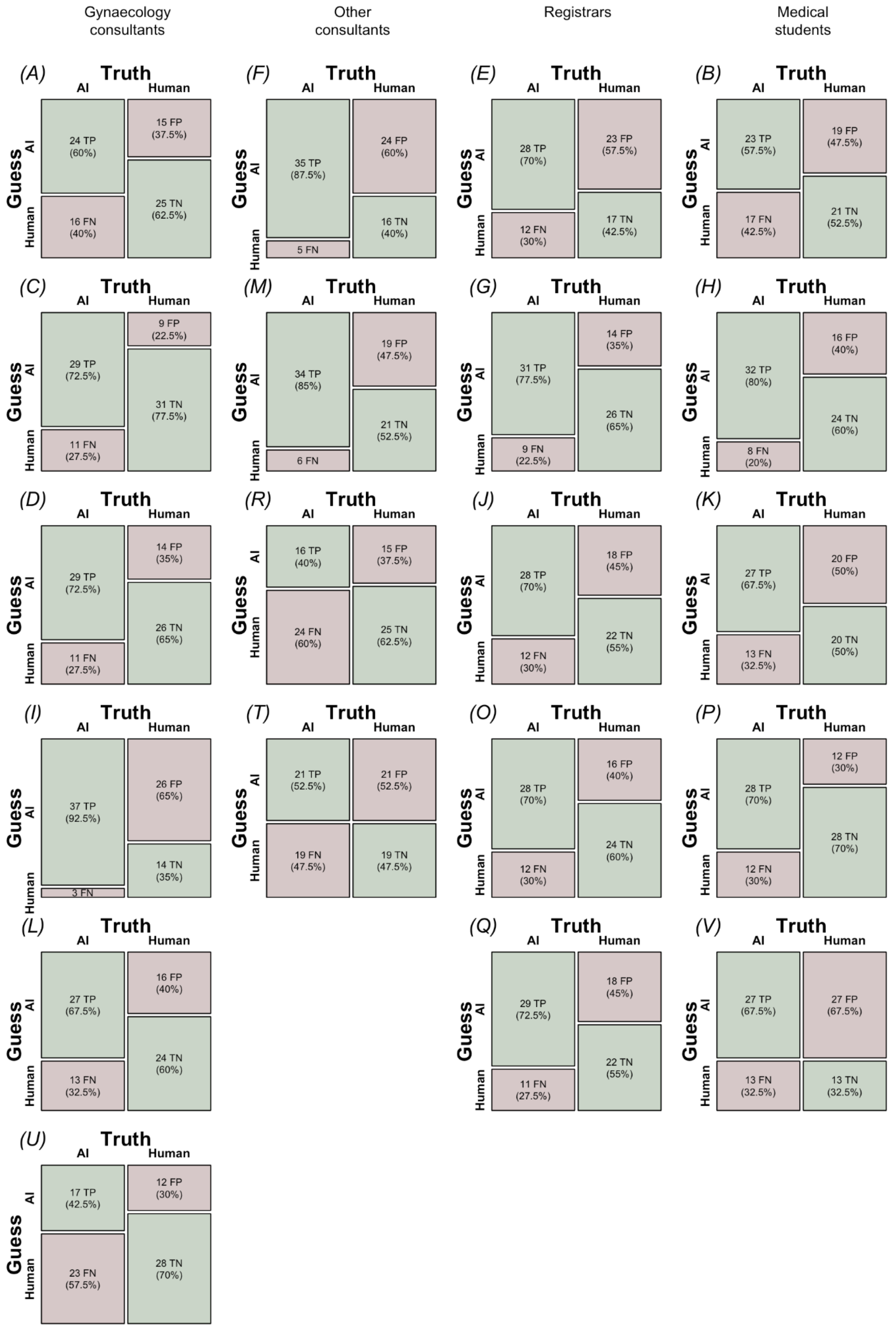

### H. Confusion matrices of accuracy of interrogator assessment for PD VTT by assessor

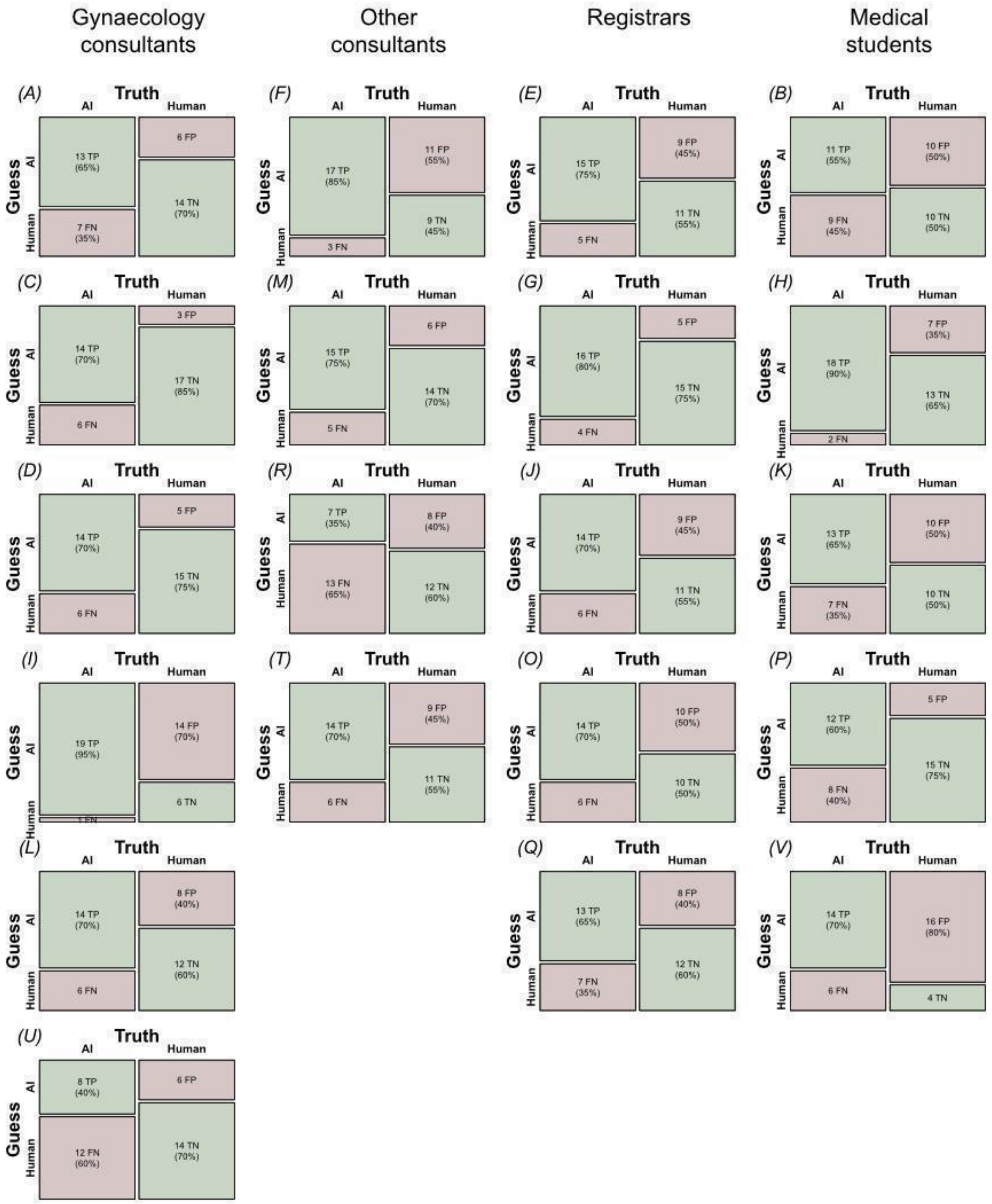

### I. Confusion matrices of accuracy of interrogator assessment for OD VTT by assessor

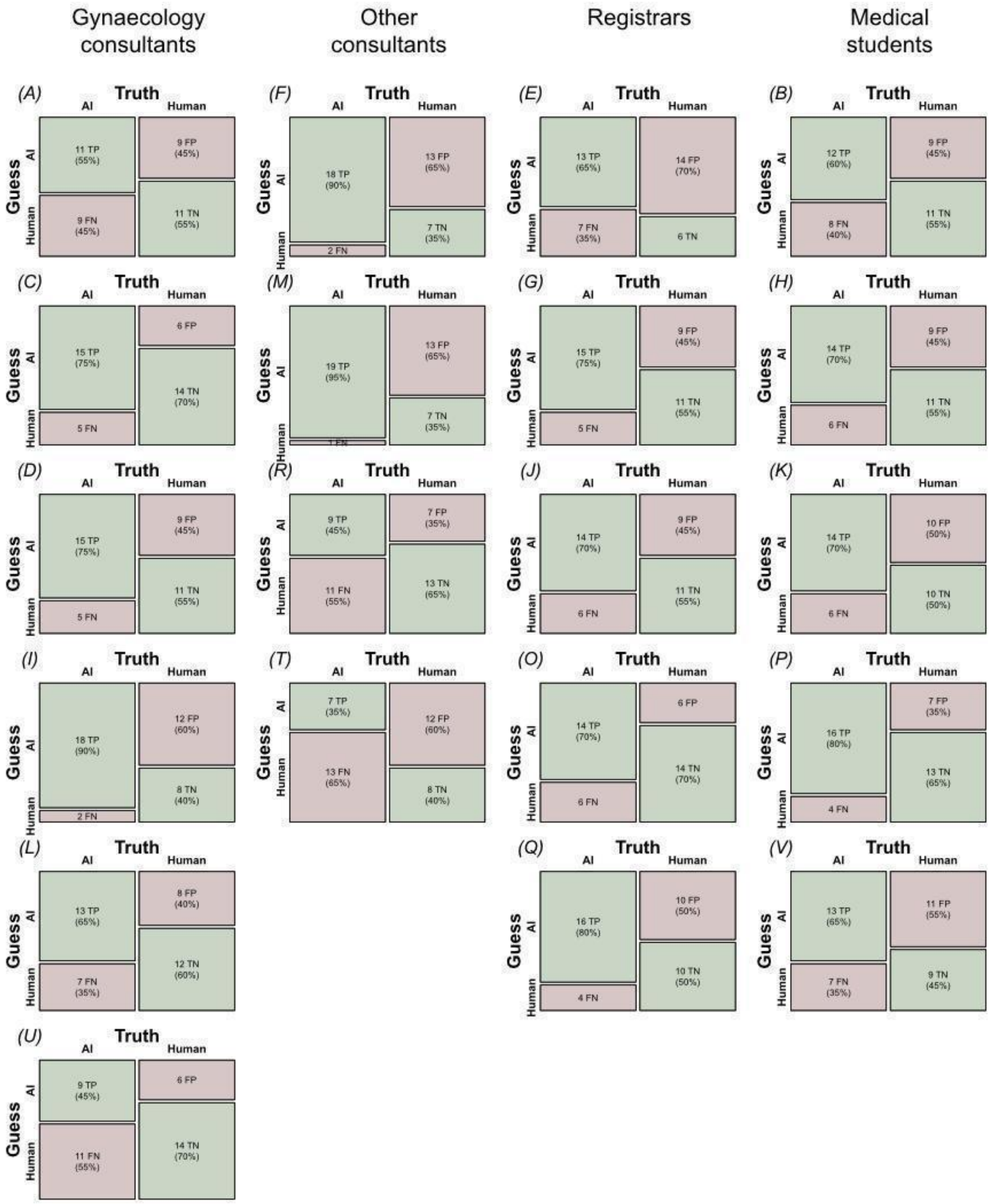
